# Use and appropriateness of Reporting Guidelines in physical therapy research: a protocol for a meta-research study

**DOI:** 10.1101/2020.05.20.20106450

**Authors:** Tiziano Innocenti, Stefano Salvioli, Silvia Giagio, Daniel Feller, Alessandro Chiarotto, Nino Cartabellotta

**Author notes:** **Corresponding author:** Tiziano Innocenti.

## Abstract

**Introduction:** Problems about reporting of a scientific study can affect dissemination of research in different manners. It is known that study methods are frequently not described in adequate detail and that results are presented ambiguously or selectively. To overcome these problems, reporting guidelines (e.g. CONSORT, PRISMA, STARD and STROBE statements) have been developed to increase the transparency of research. While recent evidence suggested that these guidelines are frequently used inappropriately in studies published in the major medical journals, the extent and appropriateness of use in physical therapy research has not been systematically evaluated. The aim of this meta-research study will be to evaluate the frequency of using reporting guidelines and their appropriate use in physical therapy published research.

**Methods and Analysis:** A cross-sectional analysis is planned on a random sample of 200 studies published in the last ten years (2010-2019) in the five generic rehabilitation journals with the highest 5-year impact factor. Randomization will be stratified for publication date to include an equal number of studies from 2010 to 2014 (n=100), and from 2015 to 2019 (n=100). Randomised controlled trials, systematic reviews, diagnostic accuracy studies and observational studies will be included. Crude prevalence estimates with 95% confidence intervals of using and appropriateness of using will be estimated. A bivariate analysis will assess the relationship between the use of reporting guidelines and the year of publication.

**Ethics and Dissemination:** Several studies have shown the positive influence of reporting guidelines on the completeness of research reporting but no one investigated the use and the appropriateness of reporting guidelines in physical therapy research. Therefore, this study will add relevant knowledge that may contribute to improve further the reporting of rehabilitation research. The results of this research will be published in a peer-reviewed journal and will be presented at relevant (inter)national scientific events.

## INTRODUCTION

Research in health sciences should have the potential to advance scientific understanding, or to improve the treatment or prevention of disease. Clinical practice and public health policy decisions should be based on high-quality research findings[1]. The purpose of a research report (e.g. a scientific article) is to communicate the design, execution, and findings of a study with precision and accuracy[2]; this is relevant for several stakeholder groups, including researchers, clinicians, systematic reviewers and patients[3]. Research reports should include all relevant information about methods and results to be useful to all these categories of potential readers. Additionally, accurate reporting of a study is essential to judge its validity and the clinical applicability of its findings[1].

Problems about reporting of a scientific study can affect research in different ways. For example, it is known that study methods are frequently not described in adequate detail and that results are presented ambiguously, incompletely or selectively[3]. The consequence is that many reports cannot be used for replication studies, or they are even harmful, as well as a waste of resources[4]. To overcome these problems, reporting guidelines (RGs) have been developed to support authors in reporting research methods and results. These are typically presented as checklists or flow diagrams that lay out the core reporting criteria required to provide a clear representation of each part of a study report (abstract, introduction, methods, results, discussion and conclusion)[5]. RGs specify a minimum set of information needed for a clear and complete account of what was done and what was found during a research study, particularly reflecting aspects that might have introduced bias into the research[1]. Nevertheless, it is important to distinguish RGs from critical appraisal tools as the objective of RGs is not to focus on the assessment of internal validity of a study. In fact, RGs provide guidance for writing an accurate research report, whereas critical appraisal tools (e.g. the Cochrane Risk of Bias 2.0 for randomized controlled trials[6]) are used to evaluate the methodological quality and/or risk of bias of a study.

RGs have been developed for a lot of different types of studies. For example, the Consolidated Standards of Reporting Trials (CONSORT Statement) is for randomized controlled trials[7, 8], the Preferred Reporting Items for Systematic Reviews and Meta-Analyses (PRISMA) Statement is for systematic reviews[9], the Strengthening the Reporting of Observational Studies in Epidemiology (STROBE) Statement is for observational designs[10], and the Standards for the Reporting of Diagnostic Accuracy Studies (STARD) Statement is for diagnostic accuracy studies[11, 12]. Many studies have confirmed that the use of RGs can improve the completeness and transparency of publications in different medical research areas[13, 14]. However, Caulley et al., in a recent cross-sectional survey[15], found that RGs of health research studies published in the major medical journals are frequently used inappropriately; for example, some authors use the RGs as a tool to guide the design and research methodology rather than an instrument to write the research report.

Research on physical therapy (PT) is a growing field in health sciences[16]. In 2014, twenty-eight rehabilitation journals have simultaneously published an editorial to highlight the need of using RGs to ensure the quality of PT and rehabilitation research[5]. Chan and colleagues concluded the editorial hoping “…that simultaneous implementation of this new reporting requirement will send a strong message to all disability and rehabilitation researchers about the need to adhere to the highest standards when performing and disseminating research.” Despite major journals in the rehabilitation field have now made RGs as mandatory item in submission systems, up to now, the extent and appropriateness of use of RGs in the PT field has not been systematically evaluated.

### Objectives

The primary objective of this meta-research study is to evaluate the frequency of using RGs and their appropriate use in PT published research.

The secondary objective of this cross-sectional analysis is to investigate the context and circumstances of using RGs by exploring the influence of year of publication and the using distribution in main field of PT (e.g. musculoskeletal, neurological, pediatrics, pelvic-floor dysfunction, and pulmonary).

## METHODS AND ANALYSIS

A cross-sectional analysis was designed in a random sample of 200 studies published in the last ten years (2010-2019) in the five generic rehabilitation journals with the highest 5-year impact factor (5-year IF) according to 2018 InCites Journal Citation Report[17]. Generic rehabilitation journals have a broad scope and can publish article in any field of PT research. The five selected journals were:

- Journal of Physiotherapy (5-year IF=6.380)
- Archives of Physical Medicine and Rehabilitation (5-year IF=3.618)
- Physical Therapy (5-year IF=3.599)
- Clinical Rehabilitation (5-year IF=3.196)
- Physiotherapy (5-year IF=3.103)

### Study selection criteria

Research reports published in the last ten years as full-text scientific articles in the aforementioned journals will be included. Articles will have to be identified in the title or in the abstract by authors as:

- Randomised controlled trial (RCT);
- Systematic review, with or without meta-analysis;
- Diagnostic accuracy study (defined also cross-sectional diagnostic study);
- Observational study (cohort, case-control and cross-sectional ones).

Unidentified reports will be labelled in one of these four categories in line with the methodology used in the study, according to the following reporting guidelines:

- CONSORT for RCTs[7, 8];
- PRISMA for systematic reviews[9];
- STARD for diagnostic studies[11, 12];
- STROBE statements and their extension for observational studies[10].

### Study selection process

An experienced librarian helped in developing the search strategy for this study. Journal tags for the five journals will be identified in Medline and a detailed search strategy will be created to find all relevant studies published in the last ten years in this database (See Table 1).

**Table 1:** Search strategy in Medline.

| # | Keywords/Tags | Detail |
| --- | --- | --- |
| 1 | "Physiotherapy"[Journal] | Journal tags in Medline |
| 2 | "Clinical rehabilitation"[Journal] |  |
| 3 | "Physical therapy"[Journal] |  |
| 4 | "Archives of physical medicine and rehabilitation"[Journal] |  |
| 5 | "Journal of physiotherapy"[Journal] |  |
| 6 | ("2010/01/01"[Date - Publication]<br>"2019/12/31"[Date - Publication]) | Searching date range |
| **Rational:** |  |  |
| (1 OR 2 OR 3 OR 4 OR 5) AND 6 |  |  |
| **Complete Search Strategy** | ((((("Physiotherapy"[Journal]) OR "Clinical rehabilitation"[Journal]) OR "Physical therapy"[Journal]) OR ("Archives of physical medicine and rehabilitation"[Journal]))) OR "Journal of physiotherapy"[Journal]) AND (("2010/01/01"[Date - Publication] : "2019/12/31"[Date - Publication])) |  |

Titles and abstracts of articles published in the years ranging from 2010 to 2019 will be screened for eligibility. Two reviewers will run the search strategy and will select all relevant articles based on title/abstract, independently. Any disagreements about inclusion of a citing article will be resolved by a third reviewer. Subsequently, the study sample of 200 hits will be selected using a computerized random sequence generator[18]. Randomization will be stratified for publication date to include an equal number of studies from 2010 to 2014 (n=100) and from 2015 to 2019 (n=100).

Study selection process will be summarized in a tailored flow diagram.

### Data collection and extraction

Endnote[19] will be used to collect and manage the study sample. Data extraction will be performed by two reviewers, independently; a third researcher will solve disagreements. A data extraction form will be used by two reviewers and the following data will be extracted:

- First author and year of publication;
- Journal;
- Study type (i.e. RCT, systematic review, diagnostic study, observational study);
- PT research field (e.g. musculoskeletal, neurological, pediatrics, pelvic-floor disfunction, cardio-pulmonary, other)
- Authors’ declaration of using a RG (yes/no and which one);
- The appropriate use of RG (yes/no).

Classification will be based on direct interpretation from the text of stated use of reporting guidelines and, when available, supplementary materials stating use of reporting guideline checklists. Use RGs as a guide to report details of the study design and results (that is, what was done and what was found) will categorize as “appropriate” use. Examples of appropriate use of these reporting guidelines are provided in Table 2. Use RGs as quality assessment checklist (e.g. risk of bias evaluation) or guidance to develop study methodolody will categorize as “inappropriate”. If the purpose of use will not state clearly, it will categorize as “unclear”.

**Table 2:**
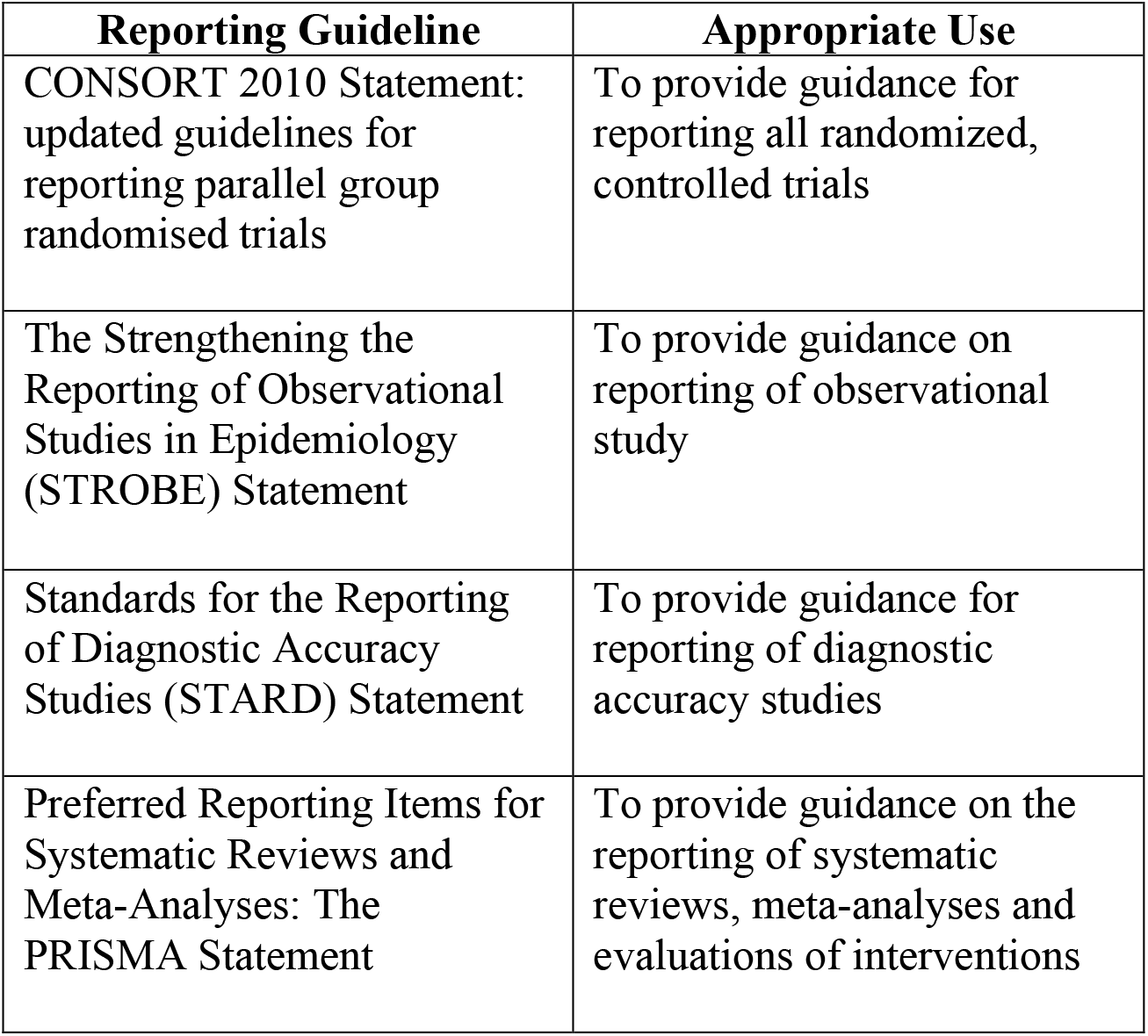
Examples of appropriate use of RG.

| **Reporting Guideline** | **Appropriate Use** |
| --- | --- |
| CONSORT 2010 Statement: updated guidelines for reporting parallel group randomised trials | To provide guidance for reporting all randomized, controlled trials |
| The Strengthening the Reporting of Observational Studies in Epidemiology (STROBE) Statement | To provide guidance on reporting of observational study |
| Standards for the Reporting of Diagnostic Accuracy Studies (STARD) Statement | To provide guidance for reporting of diagnostic accuracy studies |
| Preferred Reporting Items for Systematic Reviews and Meta-Analyses: The PRISMA Statement | To provide guidance on the reporting of systematic reviews, meta-analyses and evaluations of interventions |

### Data analysis

The primary analysis will address:

- Crude prevalence estimates with 95% confidence intervals (95% CI)[20] of articles in which authors declare the use of a RG
- Crude prevalence estimates presented alongside their 95% CI of appropriateness of use of RGs.

Secondary analysis:

- The impact of time elapsed since publication of the articles and use of RGs will be evaluated. A logistic regression analysis will be performed to assess the relationship between the use of RGs and the year of publication, with use of RG (yes/no) as dependent variable and publication year (2010-2019) as independent variable.
- We will perform in addition two subgroup analysis:
  1. Articles will be classified according to the PT research field: musculoskeletal, neurological, pediatrics, pelvic-floor disfunction, cardio-pulmonary, or other. Prevalence estimates (presented alongside 95% CIs) of articles that use a RG and their appropriateness will be calculated for each subgroup.
  2. Article will be classified according to study design: RCT, systematic review, diagnostic test accuracy, observational. We will calculate prevalence estimates (alongiside 95% CIs) of articles that use a reporting guideline and the appropriateness will be estimated for each of these groups.

## ETHICS AND DISSEMINATION

Several studies have shown the positive influence of RGs on the completeness of research reporting[21]. The adherence to these guidelines can result in a published article that contains precise information and that can better allow readers to make informed judgments. By increasing the likelihood that critical information is included in the submitted manuscript, the work of editors and external reviewers can also become more efficient[2].

Since the publication of the multi-journal editorial of Chan and colleagues[5], no study investigated the use and the appropriateness of RGs in PT research.

Therefore, this study will add relevant knowledge that may contribute to improve further the value of rehabilitation research.

This study will also align with the mission statement of the EQUATOR (Enhancing the QUAlity and Transparency of health Research) Network[22], an international collaboration aiming to enhance the reliability of medical research literature by promoting transparent, accurate reporting of research studies.. A manuscript with results will be prepared and submitted for journal publication upon project completion. The findings of the study will be disseminated at a relevant (inter)national conference. The results of this research will be published in a relevant journal in the rehabilitation category, which has peer review and qualifies physical therapy research and practice. All results of this meta-research study will also be announced at (inter)national scientific events in the area of rehabilitation and research method.

## Data Availability

All data are presented in manuscript

## AUTHORS’ CONTRIBUTIONS

TI, SS, and SG conceived and designed the study protocol. TI, SS, SG and DF designed the draft search strategy. TI, SS, SG, DF and AC were involved in conceptualising the study objectives, providing input into the search strategy, study selection criteria and plans for data extraction, with NC providing feedback. All the authors, including TI, SS, SG, DF, AC and NC, approved the final version of the manuscript.

## FUNDING STATEMENT

This research received no specific grant from any funding agency in the public, commercial or not-for-profit sectors.

## COMPETING INTERESTS

None declared.

## Notes

### Competing Interest Statement

The authors have declared no competing interest.

### Clinical Protocols

https://doi.org/10.17605/OSF.IO/ST7H8

### Funding Statement

No funding

